# Inequalities in relative cancer survival by race, immigration status, income, and education for 22 cancer sites in Canada, a cohort study

**DOI:** 10.1101/2024.05.31.24307976

**Authors:** Talía Malagón, Sarah Botting-Provost, Alissa Moore, Mariam El-Zein, Eduardo L. Franco

## Abstract

**Introduction:** There is a paucity of disaggregated data to monitor cancer health inequalities in Canada. We used data linkage to estimate site-specific cancer relative survival by race, immigration status, household income, and education level in Canada.

**Methods:** We pooled the Canadian Census Health and Environment Cohorts, which are linked datasets of 5.9 million respondents of the 2006 long-form census and 6.5 million respondents of the 2011 National Household Survey. Individual-level respondent data from these surveys were probabilistically linked with the Canadian Cancer Registry up to 2015 and with the Canadian Vital Statistics Database up to 2019. We used propensity score matching and Poisson models to calculate age-standardized relative survival by equity stratifiers for all cancers combined and for 22 individual cancer sites for the period 2006-2019.

**Results:** There were 757,485 primary cancer cases diagnosed over follow-up included in survival analyses; the age-standardized period relative survival was 72.5% at 5 years post-diagnosis. Relative survival was higher in immigrants (74.6%, 95%CI 74.3-74.8) than in Canadian-born persons (70.4%, 95%CI 70.2-70.6), and higher in racial groups with high proportions of immigrants. There was a marked social gradient by household income and education level, with 11-12% lower relative survival in cancer patients in the lowest household income and education levels than in the highest levels. Socioeconomic gradients were observed for most cancer sites, though the magnitude varied.

**Conclusions:** Despite the availability of universal healthcare in Canada, the observed differences in relative survival suggest there remain important inequities in cancer control and care.

## Introduction

There is a paucity of data to monitor and address cancer health inequalities in Canada.^1^ While net cancer survival has markedly improved over the past few decades,^2^ it is unclear whether all Canadians have benefited equally from these improvements. Data from other countries have documented significant inequalities in cancer survival by social equity stratifiers such as race, ethnicity, education, income, and immigration status.^3,4^ Some studies have examined survival by these health equity stratifiers in Canada, but they have generally been restricted to analyses from individual provinces (mostly Ontario).^5–9^ It has proved more challenging to provide pan-Canadian-level statistics because the Canadian Cancer Registry (CCR) does not collect unique health identifiers or sociodemographic data beyond age, sex, and postal code. National-level statistics on cancer survival inequalities have, therefore, mostly focused on location-based inequalities such as survival by province, rurality, and ecological neighborhood-level socioeconomic status based on residential postal code,^10,11^ rather than individual-level equity stratifiers.

Cancer survival is generally reported by cancer registries using relative survival as the estimate of net survival to cancer.^12,13^ Relative survival is calculated by dividing the observed survival in cancer cases by the expected survival for the general population, generally estimated using population life tables. Relative survival is interpretable as the expected probability of cancer survival in the absence of other causes of death. Part of the difficulty in estimating cancer survival disaggregated by health equity stratifiers comes from a lack of background mortality data to calculate expected survival. For example, relative survival by race in the US can be estimated due to the availability of life tables by race.^14^ Canadian life tables are only stratified by age, sex, and province.^15^ Because the background risk of non-cancer mortality differs by socioeconomic and demographic factors, it is not possible to calculate relative survival with life tables unless sociodemographic data can be linked to both cancer registries and vital statistics databases.^12^ The validity of relative survival is also predicated on the assumption that life tables provide a good estimate of the expected mortality risk that a person with cancer would have experienced had they not had a cancer diagnosis (the counterfactual).^13^ This assumption is questionable, as cancer cases likely differ from the general population on a number of social determinants affecting both cancer and non-cancer mortality. Methods to estimate background mortality based on more variables than those typically included in life tables could potentially lead to improved estimates of relative survival, especially when comparing groups with different background mortality rates.

Recent probabilistic data linkages since 2019 of the CCR and vital statistics with survey data by Statistics Canada make it now possible to examine health inequalities by individual-level stratifiers in cancer relative survival at the national level. Our primary objective was to estimate relative cancer survival in Canada stratified by race, income, education, immigration status, and cancer site in a representative sample to assess the presence of inequalities for health equity monitoring at the pan-Canadian level. Our secondary objective was to compare two methods for estimating the expected background mortality in different groups and assess its impact on relative survival estimates: matching of cancer cases to controls versus group-specific life tables.

## Methods

### Data sources

The CCR collects data on all new primary cancer cases diagnosed among Canadian residents since 1992. It covers all provinces and territories; however, cancer cases from the province of Québec are missing after 2010, as at the time of data linkage the province of Québec had not submitted data to the CCR since 2010. The Canadian Vital Statistics Death (CVSD) database collects information on all deaths in Canada. We used the CCR to identify primary cancer diagnoses and the CVSD to identify their date of death.

The Canadian long-form census is a mandatory survey conducted every five years targeting a representative sample of 20% of the population residing in Canada. It collects information on the demographic, social, and economic characteristics of households to support planning and government activities. In 2011, the long-form was replaced with the voluntary National Household Survey (NHS), which targeted the same population and collected the same information. While the voluntary NHS had a lower response rate (68.6%) than the mandatory 2006 long-form census (93.8%), measures were deployed to offset data quality risks and validate the representativity of results using other data sources.^16^ We used the self-reported data from these surveys to define health equity stratifiers.

The Canadian Census Health and Environment Cohorts (CanCHECs) 2006 and 2011 are a probabilistic linkage of the respondents of the 2006 long-form census and 2011 NHS with the CCR and CVSD.^17^ The linkage rates were 90.8% and 96.7% for 2006 and 2011 respondents, corresponding to 5.9 and 6.5 million individuals, respectively. Data were linked to the CCR up to 2015, and to the CVSD up to 2019. We pooled both cohorts to increase sample size and follow-up time.

### Health equity stratifiers

Health equity stratifiers were based on respondent data from the long-form and NHS. Race is a social construct based on perceived differences in physical appearance, and was categorized into the following groups based on Canadian Institute for Health Information recommendations:^18^ Black, East Asian, Indigenous, Latin American, Middle Eastern, South Asian, Southeast Asian, White, and Other. The Other group includes those who do not identify with any of the named groups or who identify as a mix of different groups. Income was measured as the total after-tax income from all household members, based on either linkage to tax files or self-report in the census questionnaire. Household income was adjusted by an equivalency factor which accounts for household size, and categorized into quintiles at the national level. Education level was measured according to a person’s highest completed degree. A Canadian-born person is someone born in Canada and a citizen by birth. An immigrant is a person who has been granted the right to live permanently in Canada by immigration authorities, including both naturalized citizens and permanent residents. A non-permanent resident is a person who has a temporary work/study permit or is a refugee claimant.

### Cancer case inclusions

We used a period survival analysis approach,^19^ including both incident cases diagnosed during follow-up (2006-2019) and prevalent cases who were within 10 years of their diagnosis on census day (mid-May 2006 or 2011). We included primary cancers among individuals aged 15 to 99 years at diagnosis; this age range was selected for consistency with national cancer statistics.^20^ We excluded cases whose diagnosis was established by autopsy only or death certificate only, cases with a missing diagnosis date, and cases whose death date preceded their diagnosis date. We allowed multiple primary cancers per person to contribute to analyses if they were diagnosed in different years. Cases were classified by cancer site using the grouping definitions of the 2021 Canadian Cancer Statistics (Supplementary Methods & Figures).^20^

Follow-up started either on the date of cancer diagnosis for incident cases, or on census day for prevalent cases. Cases contributed person-time to interval-specific conditional survival analyses for each year they were alive during follow-up, up to 10 years after their cancer diagnosis or the last date of data linkage on December 31^st^ 2019. We excluded prevalent cancer cases for the analyses stratified by income to prevent reverse causality, as a cancer diagnosis can potentially affect a household’s income.

### Statistical analyses

#### Relative survival based on matched controls

In our main analysis, we estimated relative survival using matching of cancer cases to controls to estimate the expected background mortality risk. Controls were selected among CanCHEC respondents who had no prior record of a site-specific cancer diagnosis in the CCR since 1992, and who were alive during the year of the diagnosis of their matched cancer case. Controls were matched to cancer cases using propensity score matching, using greedy nearest neighbor matching without replacement.^21^ The propensity score models included age, sex, race, household after-tax income quintile, education level, immigration status, neighborhood income level quintile, province of residence, rurality of residence, and calendar year as predictors of cancer diagnosis. Exact matches were requested for age, sex, race, and calendar year. Separate propensity score models were fit for each cancer site. Up to 3 controls were selected for each cancer case, except for the model for all cancer sites combined, where only 1 control was selected per case to reduce computational burden. The index date of start of follow-up for controls was the date of start of follow-up of their matched cancer case. We excluded controls whose date of death preceded the index date. Estimates were weighted using propensity matching weights.

We calculated relative cancer survival for each year up to 10 years post cancer diagnosis using a Poisson regression model. Dickman *et al*. (2004) proposed that relative survival could be estimated assuming a Poisson process for the number of deaths by interval since diagnosis, which assumes a piecewise constant mortality hazard function.^22^ We modified the parameterization of their proposed model to fit a Poisson model estimating the number of deaths from all causes (*µ_i_*) for observation *i*:

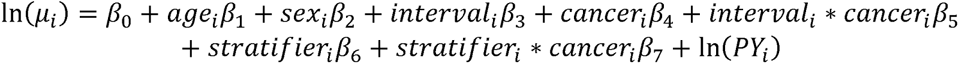

where *age_i_* is a categorical variable for age at diagnosis (15-44, 45-54, 55-64, 65-74, 75-99 years), *sex_i_* is a categorical variable for sex, *interval_i_* is a categorical variable indicating the time since cancer diagnosis in 1-year intervals, *cancer_i_* indicates whether *i* is a cancer case or a control, *stratifier_i_* is a health equity stratifier of interest, and *PY_i_* are person-years at risk during the follow-up interval.

This model estimates the excess mortality rate in cancer cases compared to controls by time interval since diagnosis (parameters *β*_4_, *β*_5_), and allows different groups to have different excess cancer mortality rates (*β*_7_) while controlling for differences in background mortality from other causes (*β*_6_). We used a type 3 test of parameters to assess whether the excess cancer mortality rate differs across groups. The model parameters were used to estimate mortality rate ratios using the exponent of model parameters, and the cumulative survival by time *t* (*S_t,i_*) in cases and controls using the transformation of the hazard approach, where *X_i_*′*β* is the vector of model covariates:

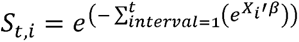

Cancer relative survival (*RS_t,i_*) was then calculated by dividing the expected survival for cancer cases by the expected survival for controls of the same age, sex, and stratifier group. Confidence intervals (95%CI) for relative survival were calculated using the log-log transformation and the delta method with the R *msm* package.^23,24^

#### Relative survival based on life tables

In secondary analyses, we recalculated relative survival using group-specific life tables stratified by race and household income quintile. The objective of this secondary analysis was to obtain relative survival estimates comparable to those reported by the Canadian Cancer Statistics for validation of sample representativity, and to compare results with relative survival estimated with the aforementioned matching methods. We calculated age, sex, income- and race-specific life tables for the entire CanCHEC 2006 & 2011 cohorts. We used the hazard transformation approach to estimate relative cancer survival using the period method.^25^ Estimates were weighted using CanCHEC survey weights. CIs for relative survival estimates were obtained through bootstrapping, using the 2.5^th^ to 97.5^th^ percentiles of 500 CanCHEC-specific bootstrapping weights. The Supplementary Methods & Figures provide more details on the methods used to generate life tables, relative survival estimates, and background mortality rates.

#### Age standardization

Relative survival estimates were age-standardized using the Canadian Cancer Survival Standard age weights for individual cancer sites.^2,20^ For all cancers combined, we used the international single standard of the EUROCARE-2 study.^26^ For Poisson models, we specified age weights in estimating equations to estimate age-standardized survival. For survival calculated with life tables, we used direct standardization.

#### Validation of estimates

To assess whether our survival estimates are representative of the Canadian population, we compared the relative survival estimates based on life tables in CanCHECs with official statistics of relative cancer survival based on life tables for all of Canada. Cancer survival has significantly improved in Canada over the past 20 years.^2^ Because our survival estimates span the period from 2006 to 2019, we compared our results to national relative survival estimates for 2006-2008 and 2015-2017 to define the target range.^20,27^

#### Ethics and confidentiality

We obtained ethical approval from the McGill University Institutional Review Board for this analysis of secondary data. To protect respondent confidentiality, analysis outputs were vetted using rules developed by Statistics Canada, which include rounding all counts to base 5 and not disclosing statistics for groups with less than 5 contributing events.

## Results

### Baseline characteristics of cancer cases and controls

The demographic characteristics of the CanCHECs have previously been described;^17,28^ we focus here on describing the characteristics of cancer cases contributing to analyses (Table 1). There were 757,485 cancer cases and 754,495 matched controls eligible for the all cancer sites combined period survival analyses. Cancer cases had a mean age at diagnosis of 63.1 years (standard deviation 14.2), and 50% were female. The majority self-identified as White (88%) and Canadian-born (75%). Cases and matched controls were comparable in terms of age, sex, race, immigration status, province of residence, rurality, household income, neighborhood income, and education level.

**Table 1.** Baseline characteristics of cancer cases aged 15-99 years at diagnosis and matched controls for all sites combined, CanCHECs 2006 & 2011.

|  | Cancer cases | Matched controls |
| --- | --- | --- |
| Number<sup>a</sup> | **757,485** | **754,495** |
| Mean age at diagnosis (SD)<sup>b</sup> | 63.1 (14.2) | 63.1 (14.2) |
|  | (%) | (%) |
| Sex |  |  |
| Female | 49.8 | 49.9 |
| Male | 50.2 | 50.1 |
| Age group on July 1<sup>st</sup> of matching year<sup>c</sup> |  |  |
| <15 years<sup>c</sup> | <0.1 | <0.1 |
| 15-19 years | 0.2 | 0.2 |
| 20-24 years | 0.4 | 0.4 |
| 25-29 years | 0.7 | 0.7 |
| 30-34 years | 1.1 | 1.1 |
| 35-39 years | 1.7 | 1.7 |
| 40-44 years | 2.8 | 2.8 |
| 45-49 years | 4.8 | 4.8 |
| 50-54 years | 7.4 | 7.4 |
| 55-59 years | 10.2 | 10.3 |
| 60-64 years | 12.9 | 13.0 |
| 65-69 years | 14.3 | 14.3 |
| 70-74 years | 13.9 | 13.9 |
| 75-79 years | 12.7 | 12.7 |
| 80-84 years | 9.7 | 9.7 |
| 85-89 years | 5.3 | 5.3 |
| 90+ years | 2.0 | 2.0 |
| Racial group |  |  |
| White | 88.0 | 88.0 |
| Indigenous | 3.6 | 3.6 |
| East Asian | 3.0 | 3.0 |
| Southeast Asian | 1.8 | 1.8 |
| South Asian | 1.1 | 1.1 |
| Middle Eastern | 1.2 | 1.2 |
| Black | 0.6 | 0.6 |
| Latin American | 0.4 | 0.4 |
| Other | 0.3 | 0.3 |
| Immigration status |  |  |
| Canadian-born | 75.1 | 75.2 |
| Immigrants | 24.7 | 24.7 |
| Non-permanent residents | 0.2 | 0.1 |
| Province |  |  |
| Newfoundland and Labrador | 1.7 | 1.7 |
| Prince Edward Island | 0.5 | 0.4 |
| Nova Scotia | 3.7 | 3.5 |
| New Brunswick | 2.9 | 2.7 |
| Québec | 15.2 | 15.3 |
| Ontario | 43.0 | 43.9 |
| Manitoba | 4.3 | 4.0 |
| Saskatchewan | 3.5 | 3.3 |
| Alberta | 10.0 | 9.9 |
| British Columbia | 14.8 | 15.0 |
| Yukon | 0.1 | 0.1 |
| Northwest Territories | 0.2 | 0.2 |
| Nunavut | 0.1 | 0.1 |
| Residence area |  |  |
| Rural | 22.1 | 21.6 |
| Urban | 77.9 | 78.4 |
| Household after-tax income quintile,<br>adjusted for household size |  |  |
| 1 (poorest) | 17.4 | 17.4 |
| 2 | 22.0 | 22.1 |
| 3 | 20.5 | 20.5 |
| 4 | 19.6 | 19.5 |
| 5 (richest) | 20.5 | 20.5 |
| Neighborhood income quintile<sup>d</sup> |  |  |
| 1 (poorest) | 18.3 | 18.4 |
| 2 | 19.6 | 19.6 |
| 3 | 20.0 | 20.0 |
| 4 | 20.3 | 20.2 |
| 5 (richest) | 21.8 | 21.8 |
| Education level<sup>e</sup> |  |  |
| None | 27.6 | 28.4 |
| Secondary school | 22.9 | 23.3 |
| Trades or registered apprenticeship | 12.4 | 12.2 |
| College | 16.0 | 15.5 |
| University, undergraduate degree | 16.0 | 15.7 |
| University, graduate or medical degree | 5.0 | 4.8 |
| Missing (<15y on census day<sup>c</sup>) | 0.1 | 0.1 |

SD=standard deviation.
<sup>a</sup> Number of cases rounded to base 5 for confidentiality.
<sup>b</sup> For controls, age on the matched cancer diagnosis date.
<sup>c</sup> Cases and controls were matched on age on July 1<sup>st</sup> of the year of cancer diagnosis (incident cases) or the census year (prevalent cases). Some cases and controls aged 14 on census day or July 1<sup>st</sup> were eligible if the cancer diagnosis date occurred in the second half of the year when they were 15 or older. Education information was not collected for individuals who were <15 years old on census day in mid-May.
<sup>d</sup> Neighborhood income quintile was derived using residential postal code with the Postal Code Conversion File Plus (PCCF+) based on 2006 census data.
<sup>e</sup> The following responses were included in each category. None: No certificate, diploma or degree. Secondary school: High school diploma or equivalent. Trades or registered apprenticeship: Registered Apprenticeship certificate; Other trades certificate or diploma. College: College, CEGEP (junior college) or other non-university certificate or diploma from a program of 3 months to less than 1 year; a program of 1 year to 2 years; or a program of more than 2 years. University undergraduate degree: University certificate or diploma below bachelor level; Bachelor's degree; University certificate or diploma above bachelor level. University, graduate or medical degree: Degree in medicine, dentistry, veterinary medicine or optometry; Master's degree; Earned doctorate.

### Relative survival compared to matched controls

Poisson model predictions for relative cancer survival by time since diagnosis for the overall cohort of cancer cases are presented in Table 2. Mortality rates for cancer cases were highest in the year following diagnosis; the mortality rate for all cancers combined was 15.9 times higher (95%CI 15.5-16.3) in cancer cases than in controls the first year after diagnosis, and declined to 1.5 times higher (95%CI 1.4-1.5) in cancer cases than in controls by the 10^th^ year after diagnosis. The predicted age-standardized relative survival for all cancers combined was 84.1% 1 year post-diagnosis, 72.5% 5 years post-diagnosis, and 67.6% 10 years post-diagnosis.

**Table 2.**
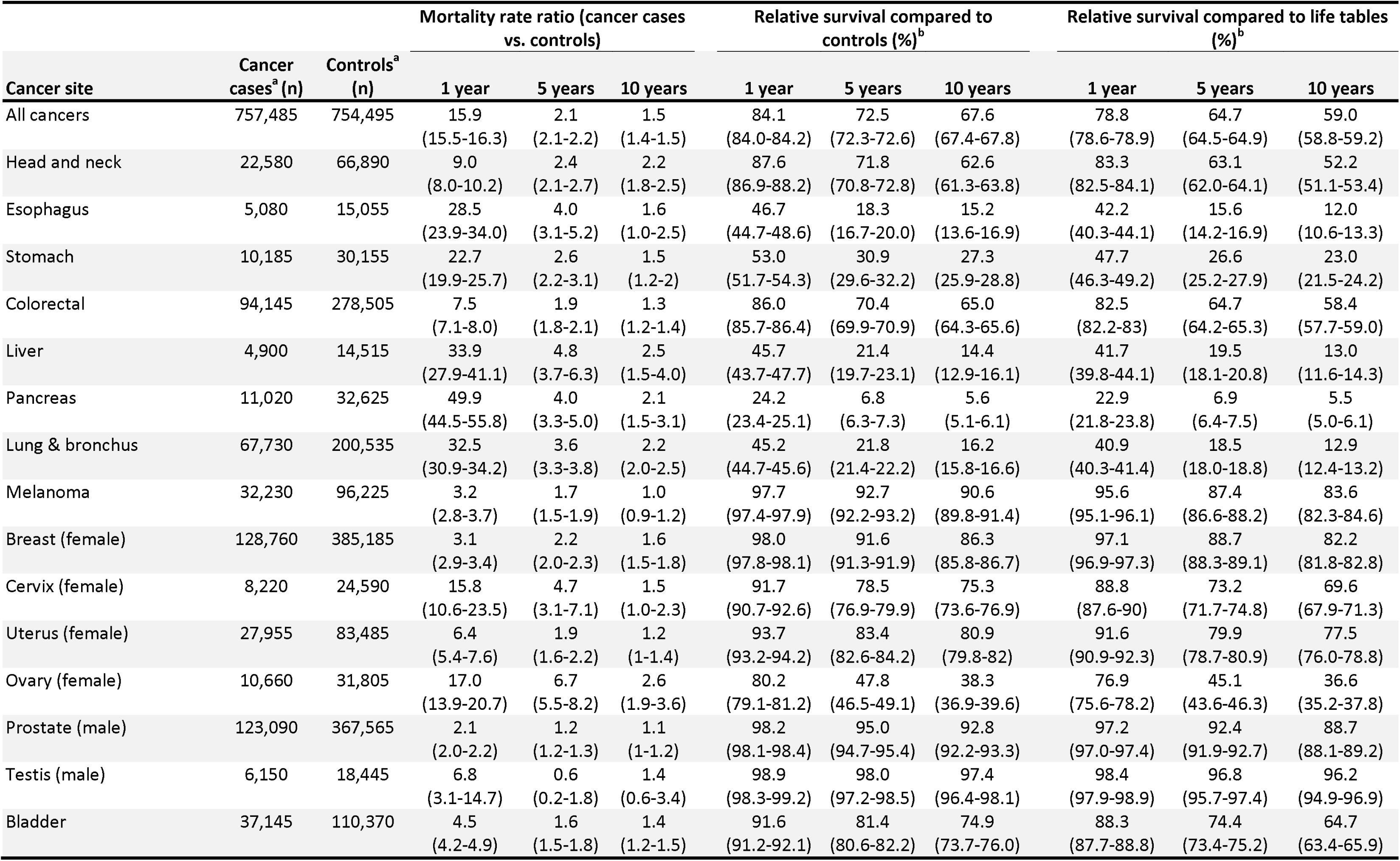
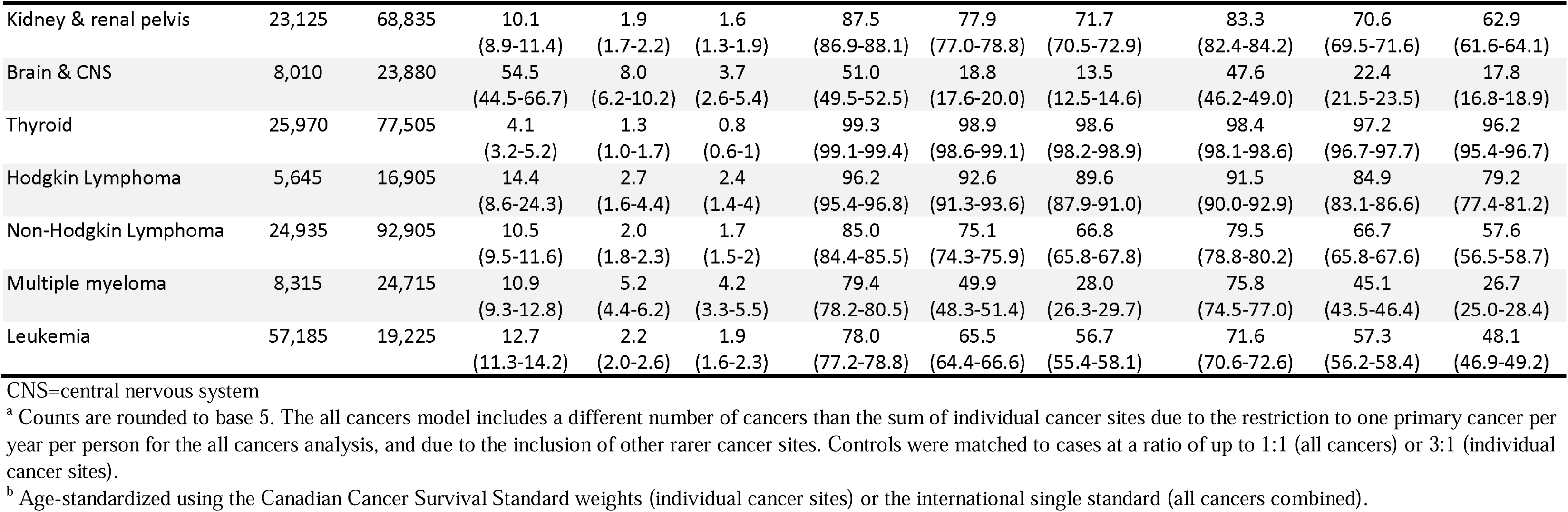
Mortality rate ratios and cumulative relative survival at 1, 5, and 10 years after cancer diagnosis, ages 15-99 years at diagnosis, estimated using two different methods. Numbers in parentheses are the 95% confidence interval.

| Cancer site | Cancer cases<sup>a</sup> (n) | Controls<sup>a</sup> (n) | Mortality rate ratio (cancer cases vs. controls) |  |  | Relative survival compared to controls (%)<sup>b</sup> |  |  | Relative survival compared to life tables (%)<sup>b</sup> |  |  |
| --- | --- | --- | --- | --- | --- | --- | --- | --- | --- | --- | --- |
|  |  |  | 1 year | 5 years | 10 years | 1 year | 5 years | 10 years | 1 year | 5 years | 10 years |
| All cancers | 757,485 | 754,495 | 15.9<br>(15.5-16.3) | 2.1<br>(2.1-2.2) | 1.5<br>(1.4-1.5) | 84.1<br>(84.0-84.2) | 72.5<br>(72.3-72.6) | 67.6<br>(67.4-67.8) | 78.8<br>(78.6-78.9) | 64.7<br>(64.5-64.9) | 59.0<br>(58.8-59.2) |
| Head and neck | 22,580 | 66,890 | 9.0<br>(8.0-10.2) | 2.4<br>(2.1-2.7) | 2.2<br>(1.8-2.5) | 87.6<br>(86.9-88.2) | 71.8<br>(70.8-72.8) | 62.6<br>(61.3-63.8) | 83.3<br>(82.5-84.1) | 63.1<br>(62.0-64.1) | 52.2<br>(51.1-53.4) |
| Esophagus | 5,080 | 15,055 | 28.5<br>(23.9-34.0) | 4.0<br>(3.1-5.2) | 1.6<br>(1.0-2.5) | 46.7<br>(44.7-48.6) | 18.3<br>(16.7-20.0) | 15.2<br>(13.6-16.9) | 42.2<br>(40.3-44.1) | 15.6<br>(14.2-16.9) | 12.0<br>(10.6-13.3) |
| Stomach | 10,185 | 30,155 | 22.7<br>(19.9-25.7) | 2.6<br>(2.2-3.1) | 1.5<br>(1.2-2) | 53.0<br>(51.7-54.3) | 30.9<br>(29.6-32.2) | 27.3<br>(25.9-28.8) | 47.7<br>(46.3-49.2) | 26.6<br>(25.2-27.9) | 23.0<br>(21.5-24.2) |
| Colorectal | 94,145 | 278,505 | 7.5<br>(7.1-8.0) | 1.9<br>(1.8-2.1) | 1.3<br>(1.2-1.4) | 86.0<br>(85.7-86.4) | 70.4<br>(69.9-70.9) | 65.0<br>(64.3-65.6) | 82.5<br>(82.2-83) | 64.7<br>(64.2-65.3) | 58.4<br>(57.7-59.0) |
| Liver | 4,900 | 14,515 | 33.9<br>(27.9-41.1) | 4.8<br>(3.7-6.3) | 2.5<br>(1.5-4.0) | 45.7<br>(43.7-47.7) | 21.4<br>(19.7-23.1) | 14.4<br>(12.9-16.1) | 41.7<br>(39.8-44.1) | 19.5<br>(18.1-20.8) | 13.0<br>(11.6-14.3) |
| Pancreas | 11,020 | 32,625 | 49.9<br>(44.5-55.8) | 4.0<br>(3.3-5.0) | 2.1<br>(1.5-3.1) | 24.2<br>(23.4-25.1) | 6.8<br>(6.3-7.3) | 5.6<br>(5.1-6.1) | 22.9<br>(21.8-23.8) | 6.9<br>(6.4-7.5) | 5.5<br>(5.0-6.1) |
| Lung & bronchus | 67,730 | 200,535 | 32.5<br>(30.9-34.2) | 3.6<br>(3.3-3.8) | 2.2<br>(2.0-2.5) | 45.2<br>(44.7-45.6) | 21.8<br>(21.4-22.2) | 16.2<br>(15.8-16.6) | 40.9<br>(40.3-41.4) | 18.5<br>(18.0-18.8) | 12.9<br>(12.4-13.2) |
| Melanoma | 32,230 | 96,225 | 3.2<br>(2.8-3.7) | 1.7<br>(1.5-1.9) | 1.0<br>(0.9-1.2) | 97.7<br>(97.4-97.9) | 92.7<br>(92.2-93.2) | 90.6<br>(89.8-91.4) | 95.6<br>(95.1-96.1) | 87.4<br>(86.6-88.2) | 83.6<br>(82.3-84.6) |
| Breast (female) | 128,760 | 385,185 | 3.1<br>(2.9-3.4) | 2.2<br>(2.0-2.3) | 1.6<br>(1.5-1.8) | 98.0<br>(97.8-98.1) | 91.6<br>(91.3-91.9) | 86.3<br>(85.8-86.7) | 97.1<br>(96.9-97.3) | 88.7<br>(88.3-89.1) | 82.2<br>(81.8-82.8) |
| Cervix (female) | 8,220 | 24,590 | 15.8<br>(10.6-23.5) | 4.7<br>(3.1-7.1) | 1.5<br>(1.0-2.3) | 91.7<br>(90.7-92.6) | 78.5<br>(76.9-79.9) | 75.3<br>(73.6-76.9) | 88.8<br>(87.6-90) | 73.2<br>(71.7-74.8) | 69.6<br>(67.9-71.3) |
| Uterus (female) | 27,955 | 83,485 | 6.4<br>(5.4-7.6) | 1.9<br>(1.6-2.2) | 1.2<br>(1-1.4) | 93.7<br>(93.2-94.2) | 83.4<br>(82.6-84.2) | 80.9<br>(79.8-82) | 91.6<br>(90.9-92.3) | 79.9<br>(78.7-80.9) | 77.5<br>(76.0-78.8) |
| Ovary (female) | 10,660 | 31,805 | 17.0<br>(13.9-20.7) | 6.7<br>(5.5-8.2) | 2.6<br>(1.9-3.6) | 80.2<br>(79.1-81.2) | 47.8<br>(46.5-49.1) | 38.3<br>(36.9-39.6) | 76.9<br>(75.6-78.2) | 45.1<br>(43.6-46.3) | 36.6<br>(35.2-37.8) |
| Prostate (male) | 123,090 | 367,565 | 2.1<br>(2.0-2.2) | 1.2<br>(1.2-1.3) | 1.1<br>(1-1.2) | 98.2<br>(98.1-98.4) | 95.0<br>(94.7-95.4) | 92.8<br>(92.2-93.3) | 97.2<br>(97.0-97.4) | 92.4<br>(91.9-92.7) | 88.7<br>(88.1-89.2) |
| Testis (male) | 6,150 | 18,445 | 6.8<br>(3.1-14.7) | 0.6<br>(0.2-1.8) | 1.4<br>(0.6-3.4) | 98.9<br>(98.3-99.2) | 98.0<br>(97.2-98.5) | 97.4<br>(96.4-98.1) | 98.4<br>(97.9-98.9) | 96.8<br>(95.7-97.4) | 96.2<br>(94.9-96.9) |
| Bladder | 37,145 | 110,370 | 4.5<br>(4.2-4.9) | 1.6<br>(1.5-1.8) | 1.4<br>(1.2-1.5) | 91.6<br>(91.2-92.1) | 81.4<br>(80.6-82.2) | 74.9<br>(73.7-76.0) | 88.3<br>(87.7-88.8) | 74.4<br>(73.4-75.2) | 64.7<br>(63.4-65.9) |
| Kidney & renal pelvis | 23,125 | 68,835 | 10.1<br>(8.9-11.4) | 1.9<br>(1.7-2.2) | 1.6<br>(1.3-1.9) | 87.5<br>(86.9-88.1) | 77.9<br>(77.0-78.8) | 71.7<br>(70.5-72.9) | 83.3<br>(82.4-84.2) | 70.6<br>(69.5-71.6) | 62.9<br>(61.6-64.1) |
| Brain & CNS | 8,010 | 23,880 | 54.5<br>(44.5-66.7) | 8.0<br>(6.2-10.2) | 3.7<br>(2.6-5.4) | 51.0<br>(49.5-52.5) | 18.8<br>(17.6-20.0) | 13.5<br>(12.5-14.6) | 47.6<br>(46.2-49.0) | 22.4<br>(21.5-23.5) | 17.8<br>(16.8-18.9) |
| Thyroid | 25,970 | 77,505 | 4.1<br>(3.2-5.2) | 1.3<br>(1.0-1.7) | 0.8<br>(0.6-1) | 99.3<br>(99.1-99.4) | 98.9<br>(98.6-99.1) | 98.6<br>(98.2-98.9) | 98.4<br>(98.1-98.6) | 97.2<br>(96.7-97.7) | 96.2<br>(95.4-96.7) |
| Hodgkin Lymphoma | 5,645 | 16,905 | 14.4<br>(8.6-24.3) | 2.7<br>(1.6-4.4) | 2.4<br>(1.4-4) | 96.2<br>(95.4-96.8) | 92.6<br>(91.3-93.6) | 89.6<br>(87.9-91.0) | 91.5<br>(90.0-92.9) | 84.9<br>(83.1-86.6) | 79.2<br>(77.4-81.2) |
| Non-Hodgkin Lymphoma | 24,935 | 92,905 | 10.5<br>(9.5-11.6) | 2.0<br>(1.8-2.3) | 1.7<br>(1.5-2) | 85.0<br>(84.4-85.5) | 75.1<br>(74.3-75.9) | 66.8<br>(65.8-67.8) | 79.5<br>(78.8-80.2) | 66.7<br>(65.8-67.6) | 57.6<br>(56.5-58.7) |
| Multiple myeloma | 8,315 | 24,715 | 10.9<br>(9.3-12.8) | 5.2<br>(4.4-6.2) | 4.2<br>(3.3-5.5) | 79.4<br>(78.2-80.5) | 49.9<br>(48.3-51.4) | 28.0<br>(26.3-29.7) | 75.8<br>(74.5-77.0) | 45.1<br>(43.5-46.4) | 26.7<br>(25.0-28.4) |
| Leukemia | 57,185 | 19,225 | 12.7<br>(11.3-14.2) | 2.2<br>(2.0-2.6) | 1.9<br>(1.6-2.3) | 78.0<br>(77.2-78.8) | 65.5<br>(64.4-66.6) | 56.7<br>(55.4-58.1) | 71.6<br>(70.6-72.6) | 57.3<br>(56.2-58.4) | 48.1<br>(46.9-49.2) |

CNS=central nervous system
<sup>a</sup> Counts are rounded to base 5. The all cancers model includes a different number of cancers than the sum of individual cancer sites due to the restriction to one primary cancer per year per person for the all cancers analysis, and due to the inclusion of other rarer cancer sites. Controls were matched to cases at a ratio of up to 1:1 (all cancers) or 3:1 (individual cancer sites).
<sup>b</sup> Age-standardized using the Canadian Cancer Survival Standard weights (individual cancer sites) or the international single standard (all cancers combined).

There were significant differences in relative survival by race, immigration status, household income, and education level for all cancers combined (Figure 1, Tables 3 & 4). Specifically, the 5-year relative cancer survival was higher in non-White and non-Indigenous racial groups (72.0-76.9%) than in White (71.6%, 95%CI 71.4-71.8) and Indigenous (58.7%, 95%CI 57.9-59.4) persons, and was higher in immigrants (74.6%, 95%CI 74.3-74.8) than in Canadian-born persons (70.4%, 95%CI 70.2-70.6) (Table 3). There was a strong gradient in relative cancer survival by household income and education level, with 5-year relative cancer survival being lowest in the poorest income quintile (63.7%, 95%CI 63.3-64.1) and highest in the richest income quintile (75.2%, 95%CI 74.9-75.6), and lowest in those with no secondary school diploma (67.7%, 95%CI 67.4-68.0) compared with those with a graduate or medical university degree (79.3%, 95%CI 78.8-79.8) (Table 4). Estimates for all 10 years of follow-up by group can be found in the Supplementary Table 1. Similar social gradients were observed across most cancer sites, though the type 3 test for heterogeneity was not always significant due to low case numbers for some cancer sites, leading to higher uncertainty in estimates.

**Figure 1.**
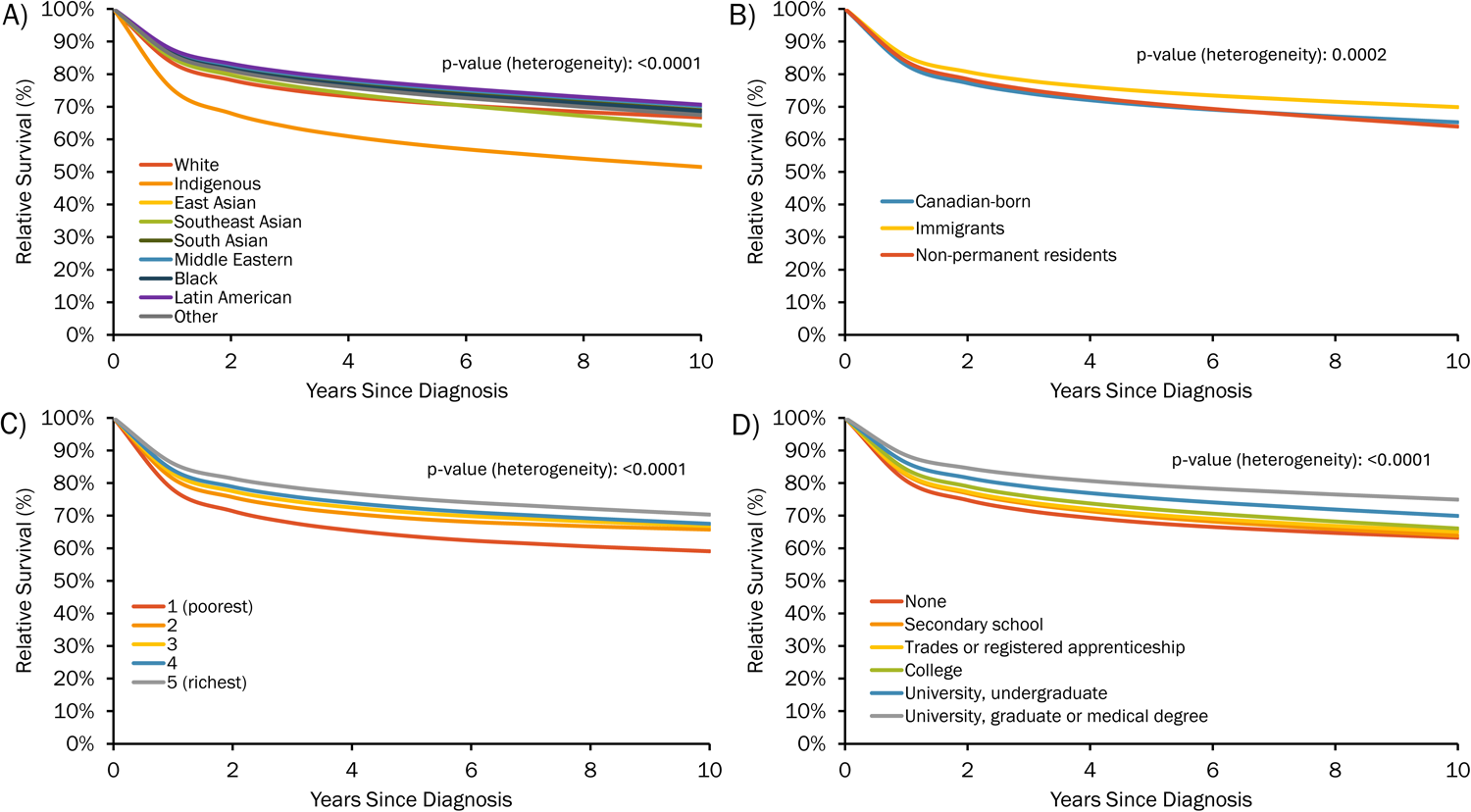
10-year relative survival for all cancers combined compared to matched controls, ages 15-99, stratified by A) race, B) immigration status, C) household income quintile, and D) education level. P-values represent a type III test for the interaction effect of each characteristic on the excess cancer mortality rate. Estimates are age-standardized using the weights of the international single standard from the EUROCARE-2 study.26

**Table 3.** Age-standardized 5-year relative survival (%) by cancer site, race, and immigration status, ages 15-99 years at diagnosis, compared with matched controls. Numbers in parentheses are the 95% confidence interval.

| Cancer site | Racial group |  |  |  |  |  |  |  |  | p-value<sup>a</sup> | Immigration status |  |  | p-value |
| --- | --- | --- | --- | --- | --- | --- | --- | --- | --- | --- | --- | --- | --- | --- |
|  | White | Indigenous | East Asian | Southeast Asian | South Asian | Middle Eastern | Black | Latin American | Other |  | Canadian-born | Immigrants | Non-permanent residents |  |
| All cancers | 71.6<br>(71.4-71.8) | 58.7<br>(57.9-59.4) | 74.2<br>(73.5-74.8) | 72.0<br>(70.8-73.1) | 75.3<br>(74.4-76.2) | 76.4<br>(74.8-77.9) | 74.9<br>(73.8-76) | 76.9<br>(74.8-78.8) | 74.2<br>(72-76.2) | <0.0001 | 70.4<br>(70.2-70.6) | 74.6<br>(74.3-74.8) | 70.9<br>(67.7-73.9) | 0.0002 |
| Head and neck | 70.9<br>(69.9-71.9) | 55.9<br>(51.7-59.8) | 73.3<br>(69.3-77.0) | 73.4<br>(65.1-80.1) | 64.9<br>(59.7-69.5) | 79.3<br>(67-87.5) | 66.7<br>(56.3-75.2) | 79.3<br>(62.9-89.1) | 73.5<br>(60.0-83.1) | <0.0001 | 69.5<br>(68.4-70.5) | 73.4<br>(71.8-74.9) | 66.4<br>(46.1-80.6) | 0.4142 |
| Esophagus | 17.3<br>(16.0-18.7) | 15.0<br>(10.4-20.4) | 15.3<br>(8.8-23.4) | 10.2<br>(1.1-31.6) | 27.7<br>(18.5-37.7) | 29.2<br>(7.9-55.1) | 9.8<br>(3-21.3) | 11.3<br>(0.8-37.5) | 29.5<br>(7.0-57.1) | 0.5687 | 16.5<br>(15.2-18.0) | 20.6<br>(18.1-23.2) | 18.1<br>(2.0-47.3) | 0.9065 |
| Stomach | 28.4<br>(27.1-29.6) | 17.6<br>(14.2-21.3) | 40.9<br>(36.5-45.2) | 37.3<br>(28.1-46.4) | 32.4<br>(24.9-40) | 46.8<br>(34.8-58) | 38.9<br>(30.3-47.4) | 35.1<br>(23.9-46.6) | 35.9<br>(21.6-50.4) | 0.004 | 26.3<br>(24.9-27.6) | 34.9<br>(33.0-36.7) | 24.0<br>(9.1-42.6) | 0.1005 |
| Colorectal | 69.5<br>(69.0-70.0) | 58.6<br>(56.5-60.6) | 73.1<br>(71.1-75.0) | 67.9<br>(63.9-71.6) | 74.2<br>(70.5-77.5) | 72.5<br>(66.7-77.5) | 67.2<br>(63.0-71.0) | 72.0<br>(64.2-78.4) | 72.6<br>(64.7-79.0) | <0.0001 | 68.0<br>(67.4-68.5) | 72.8<br>(72-73.6) | 65.7<br>(54.7-74.6) | 0.7319 |
| Liver | 18.4<br>(16.9-19.9) | 9.7<br>(7.0-13.0) | 36.8<br>(32.6-41.0) | 32.2<br>(24.9-39.7) | 26.2<br>(18.8-34.1) | 30.5<br>(15.7-46.6) | 17.3<br>(9.5-27.0) | 39.3<br>(20.7-57.5) | 33.2<br>(19.1-48.1) | 0.6506 | 15.6<br>(14.2-17.1) | 30.3<br>(27.9-32.7) | 14.4<br>(1.1-43.2) | 0.3573 |
| Pancreas | 6.3<br>(5.8-6.8) | 1.4<br>(0.9-2.1) | 11.8<br>(8.9-15.1) | 15.3<br>(9.6-22.2) | 10.7<br>(7.1-15.3) | 10.5<br>(4.3-20.1) | 9.5<br>(5.9-14.0) | 16.9<br>(8.0-28.6) | 8.5<br>(2.9-17.9) | 0.0119 | 5.6<br>(5.2-6.1) | 8.8<br>(7.9-9.8) | 15.4<br>(2.0-41.1) | 0.631 |
| Lung & bronchus | 19.6<br>(19.3-20.0) | 13.3<br>(12.1-14.5) | 28.6<br>(26.5-30.8) | 27.4<br>(23.7-31.2) | 25.4<br>(21.4-29.6) | 25.4<br>(19.3-32.0) | 19.9<br>(16.0-24.2) | 31.3<br>(22.2-40.8) | 28.8<br>(20.3-37.8) | <0.0001 | 18.5<br>(18.1-18.8) | 24.9<br>(24.1-25.6) | 30.6<br>(20.5-41.2) | 0.0018 |
| Melanoma | 92.2<br>(91.6-92.7) | 84.4<br>(76.9-89.6) | 85.9<br>(74.2-92.6) | 84.7<br>(62.8-94.2) | 75.2<br>(59.8-85.4) | 96.8<br>(63.7-99.8) | 86.7<br>(67.5-94.9) | 73.3<br>(53.6-85.7) | 94.7<br>(0.2-100.0) | 0.0334 | 92.2<br>(91.5-92.7) | 91.7<br>(90.6-92.6) | 84.9<br>(52.3-95.9) | 0.2852 |
| Breast (female) | 91.7<br>(91.4-92) | 88.7<br>(87.2-90.1) | 92.8<br>(91.7-93.7) | 90.6<br>(88.8-92.1) | 91.0<br>(89.5-92.3) | 91.3<br>(88.6-93.4) | 87.9<br>(85.7-89.8) | 89.0<br>(85.0-92.0) | 90.0<br>(86.2-92.8) | <0.0001 | 91.4<br>(91.1-91.8) | 92.1<br>(91.6-92.5) | 90.9<br>(84.7-94.6) | 0.0005 |
| Cervix (female) | 78.8<br>(77.1-80.3) | 69.5<br>(64.5-74.1) | 86.5<br>(81.2-90.4) | 79.1<br>(71.0-85.2) | 85.2<br>(78.3-90.1) | 70.5<br>(34.9-89.0) | 82.8<br>(73.6-89) | 92.6<br>(77.1-97.7) | 89.3<br>(75.5-95.6) | 0.9237 | 77.1<br>(75.4-78.7) | 83.5<br>(81.0-85.7) | 85.8<br>(56.9-96.0) | 0.5195 |
| Uterus (female) | 84.3<br>(83.4-85.1) | 74.1<br>(68.4-78.9) | 82.5<br>(78.6-85.8) | 74.5<br>(68.6-79.5) | 80.4<br>(76.2-84) | 78.5<br>(65.4-87.1) | 68.0<br>(61.2-73.9) | 82.3<br>(70.4-89.8) | 76.7<br>(63.7-85.6) | <0.0001 | 83.7<br>(82.8-84.6) | 82.6<br>(81.2-83.8) | 89.2<br>(66.7-96.8) | 0.0002 |
| Ovary (female) | 47.3<br>(45.9-48.7) | 41.2<br>(35.3-47.1) | 57.3<br>(51.0-63.1) | 61.9<br>(52.0-70.4) | 46.6<br>(40.0-52.9) | 44.5<br>(30.2-57.8) | 51.3<br>(39.9-61.5) | 55.8<br>(37.9-70.4) | 42.5<br>(26.3-57.9) | 0.0042 | 46.7<br>(45.2-48.2) | 50.7<br>(48.4-52.9) | 49.1<br>(26.6-68.2) | 0.337 |
| Prostate (male) | 97.6<br>(97.4-97.8) | 95.8<br>(94.7-96.7) | 98.8<br>(98.1-99.2) | 98.2<br>(96.9-99.0) | 97.9<br>(97.1-98.5) | 98.3<br>(96.3-99.2) | 98.1<br>(97.3-98.7) | 99.2<br>(94.9-99.9) | NE<sup>b</sup> | 0.4772 | 97.6<br>(97.3-97.7) | 97.9<br>(97.7-98.2) | 97.1<br>(88.3-99.3) | 0.4874 |
| Testis (male) | NE<sup>c</sup> | NE<sup>c</sup> | NE<sup>c</sup> | NE<sup>c</sup> | NE<sup>c</sup> | NE<sup>c</sup> | NE<sup>c</sup> | NE<sup>c</sup> | NE<sup>c</sup> | NE<sup>c</sup> | 97.9<br>(97.1-98.5) | 98.4<br>(97.0-99.1) | 97.0<br>(80.7-99.6) | 0.8324 |
| Bladder | 80.7<br>(79.9-81.5) | 67.2<br>(61.3-72.4) | 85.2<br>(81.1-88.4) | 72.6<br>(61.2-81.2) | 83.5<br>(77.6-87.9) | 83.4<br>(75.6-88.8) | 81.5<br>(72.0-88.0) | 86.4<br>(67.3-94.7) | 79.5<br>(59.6-90.3) | 0.0682 | 79.8<br>(78.9-80.6) | 83.0<br>(81.8-84.1) | 76.0<br>(47.6-90.4) | 0.4385 |
| Kidney & renal pelvis | 77.1<br>(76.1-78.0) | 68.4<br>(65.1-71.4) | 79.9<br>(75.6-83.5) | 71.9<br>(63.3-78.8) | 85.8<br>(80.7-89.7) | 88.0<br>(79.1-93.3) | 82.1<br>(74.9-87.4) | 79.1<br>(69.4-86.0) | 76.8<br>(61.1-86.8) | 0.1202 | 75.5<br>(74.5-76.4) | 80.7<br>(79.3-82) | 89.4<br>(49.9-98.2) | 0.636 |
| Brain & CNS | 17.3<br>(16.2-18.4) | 19.6<br>(14.0-25.8) | 22.2<br>(15.9-29.0) | 27.1<br>(16.3-39.2) | 29.9<br>(23.4-36.7) | 16.5<br>(8.0-27.6) | 21.8<br>(13.5-31.3) | 25.0<br>(9.5-44.1) | 14.9<br>(4.9-30.0) | 0.0796 | 16.9<br>(15.7-18.0) | 21.7<br>(19.7-23.8) | 14.7<br>(2.5-37.0) | 0.5919 |
| Thyroid | 98.7 | 95.1 | 99.1 | 99.1 | 98.8 | 99.2 | 99.1 | 99.2 | 99.2 | 0.8404 | 98.6 | 99.0 | 97.5 | 0.6864 |
|  | (98.3-98.9) | (92.3-96.9) | (98.2-99.6) | (97.4-99.7) | (97.5-99.5) | (96.0-99.9) | (97.0-99.7) | (92.2-99.9) | (93.9-99.9) |  | (98.2-98.8) | (98.6-99.2) | (90.4-99.4) |  |
| Hodgkin Lymphoma | 91.7 | 88.9 | 93.6 | 97.2 | 93.7 | 92.7 | 95.0 | 94.4 | 89.4 | 0.8654 | 91.7 | 92.7 | 77.5 | 0.6408 |
|  | (90.3-92.8) | (80.1-94.0) | (84.3-97.5) | (68.1-99.8) | (89.1-96.4) | (82.1-97.1) | (81.7-98.7) | (69.6-99.1) | (63.8-97.2) |  | (90.4-92.9) | (90.7-94.3) | (42.9-92.6) |  |
| Non-Hodgkin Lymphoma | 74.5 | 62.3 | 74.2 | 61.5 | 68.7 | 76.9 | 62.2 | 84.4 | 74.1 | <0.0001 | 73.5 | 74.7 | 80.1 | 0.0564 |
|  | (73.7-75.3) | (58.1-66.1) | (70.6-77.4) | (54.9-67.4) | (64.2-72.8) | (69.2-82.9) | (56-67.9) | (73.2-91.2) | (57.9-84.8) |  | (72.6-74.4) | (73.4-75.9) | (55.1-92.1) |  |
| Multiple myeloma | 47.3 | 37.5 | 53.6 | 65.2 | 55.5 | 49.6 | 59.9 | 65.3 | 73.4 | 0.2222 | 46.6 | 51.6 | 66.3 | 0.3127 |
|  | (45.7-48.8) | (31.4-43.6) | (45.0-61.4) | (53.7-74.6) | (48.3-62.1) | (36.5-61.3) | (52.3-66.8) | (42.8-80.8) | (53.0-86.1) |  | (44.9-48.3) | (49.2-53.9) | (30.2-86.8) |  |
| Leukemia | 65.4 | 45.3 | 51.8 | 50.2 | 66.8 | 55.2 | 51.7 | 65.8 | 69.7 | <0.0001 | 63.8 | 66.2 | 65.4 | 0.1223 |
|  | (64.3-66.4) | (39.3-51.2) | (45.6-57.6) | (39.2-60.2) | (61.1-71.9) | (44.9-64.4) | (42.6-60.0) | (49.7-77.8) | (50.3-82.7) |  | (62.6-65) | (64.4-67.8) | (45.7-79.4) |  |

CNS=central nervous system; NE=not estimable. Age-standardized using the Canadian Cancer Survival Standard weights (individual cancer sites) or the international single standard (all cancers combined).
<sup>a</sup> type III test for interaction effect between the characteristic and cancer status; tests whether there is evidence of heterogeneity in excess cancer mortality rates across groups over the entire 10-year period, not just at 5 years.
<sup>b</sup> Not estimable due to too few deaths in controls.
<sup>c</sup> The model for testicular cancer by race did not converge due to data sparsity, with too few deaths in some racial groups to estimate relative cancer survival.

**Table 4.** Age-standardized 5-year relative survival by cancer site, household income quintile, and education level, ages 15-99 years at diagnosis, compared with matched controls. Numbers in parentheses are the 95% confidence interval.

| Cancer site | Household income quintile |  |  |  |  |  | Education level |  |  |  |  |  |  |
| --- | --- | --- | --- | --- | --- | --- | --- | --- | --- | --- | --- | --- | --- |
|  | 1 (poorest) | 2 | 3 | 4 | 5 (richest) | p-value | None | Secondary school | Trades or registered apprenticeship | College | University, undergraduate | University, graduate or medical | p-value |
| All cancers | 63.7<br>(63.3-64.1) | 69.1<br>(68.7-69.4) | 70.9<br>(70.6-71.3) | 72.3<br>(71.9-72.6) | 75.2<br>(74.9-75.6) | <0.0001 | 67.7<br>(67.4-68.0) | 69.6<br>(69.3-69.9) | 70.3<br>(70.0-70.7) | 72.0<br>(71.7-72.3) | 75.3<br>(75.0-75.7) | 79.3<br>(78.8-79.8) | <0.0001 |
| Head and neck | 58.8<br>(56.4-61.1) | 68.0<br>(65.9-70.1) | 73.0<br>(70.9-75.0) | 72.7<br>(70.6-74.7) | 75.7<br>(73.7-77.6) | 0.0017 | 68.5<br>(67.0-70.0) | 68.7<br>(67.0-70.3) | 68.9<br>(66.9-70.9) | 70.6<br>(68.5-72.6) | 73.6<br>(71.5-75.6) | 78.6<br>(75.2-81.7) | <0.0001 |
| Esophagus | 12.0<br>(9.8-14.5) | 19.4<br>(16.8-22.2) | 18.9<br>(16.2-21.7) | 17.0<br>(14.6-19.6) | 20.3<br>(17.6-23.3) | <0.0001 | 17.3<br>(15.2-19.4) | 15.8<br>(13.7-18.0) | 17.0<br>(14.3-19.8) | 18.4<br>(15.6-21.3) | 17.9<br>(14.9-21.1) | 26.3<br>(19.9-33.2) | <0.0001 |
| Stomach | 26.9<br>(24.4-29.5) | 29.9<br>(27.5-32.3) | 30.4<br>(27.9-32.9) | 27.1<br>(24.6-29.6) | 29.3<br>(26.7-31.9) | <0.0001 | 29.1<br>(27.3-30.9) | 27.1<br>(25-29.3) | 30.1<br>(27.4-32.8) | 27.8<br>(25.0-30.5) | 31.6<br>(28.6-34.6) | 37.6<br>(32.2-43.0) | <0.0001 |
| Colorectal | 63.2<br>(61.9-64.5) | 68.9<br>(67.8-69.9) | 69.4<br>(68.3-70.5) | 70.2<br>(69.1-71.3) | 70.8<br>(69.6-71.9) | <0.0001 | 68.5<br>(67.8-69.3) | 67.7<br>(66.8-68.6) | 68.7<br>(67.6-69.9) | 69.1<br>(68.0-70.2) | 69.9<br>(68.8-71.0) | 72.8<br>(70.9-74.7) | <0.0001 |
| Liver | 17.4<br>(15.0-20.0) | 20.3<br>(17.6-23.2) | 23.2<br>(20.3-26.3) | 21.4<br>(18.5-24.6) | 23.7<br>(20.5-27.1) | 0.0006 | 18.9<br>(16.9-21.0) | 22.3<br>(19.7-24.9) | 19.4<br>(16.5-22.5) | 18.5<br>(15.5-21.7) | 25.4<br>(21.8-29.3) | 33.3<br>(26.2-40.4) | 0.0008 |
| Pancreas | 4.5<br>(3.8-5.4) | 5.9<br>(5.1-6.9) | 6.6<br>(5.7-7.5) | 7.7<br>(6.7-8.8) | 8.0<br>(6.9-9.1) | <0.0001 | 5.6<br>(5.0-6.3) | 5.4<br>(4.7-6.2) | 7.1<br>(6.0-8.3) | 7.1<br>(6.1-8.2) | 8.3<br>(7.1-9.6) | 6.9<br>(5.3-8.9) | <0.0001 |
| Lung & bronchus | 15.4<br>(14.8-16.1) | 19.9<br>(19.2-20.6) | 20.6<br>(19.9-21.4) | 21.1<br>(20.2-21.9) | 21.7<br>(20.8-22.7) | <0.0001 | 18.7<br>(18.2-19.2) | 18.9<br>(18.2-19.5) | 19.8<br>(19.0-20.7) | 20.1<br>(19.2-21.0) | 23.6<br>(22.5-24.8) | 27.8<br>(25.4-30.3) | <0.0001 |
| Melanoma | 89.6<br>(87.3-91.5) | 92.2<br>(90.7-93.4) | 92.0<br>(90.6-93.1) | 92.5<br>(91.2-93.6) | 92.4<br>(91.4-93.3) | 0.008 | 90.8<br>(89.6-91.9) | 90.9<br>(89.8-91.8) | 92.4<br>(91.1-93.6) | 92.1<br>(91.0-93.1) | 92.2<br>(91.2-93.1) | 93.5<br>(92.0-94.8) | 0.0029 |
| Breast (female) | 89.7<br>(88.8-90.5) | 91.6<br>(90.9-92.2) | 92.0<br>(91.4-92.7) | 92.4<br>(91.7-93.0) | 92.3<br>(91.7-92.9) | <0.0001 | 91.7<br>(91.1-92.2) | 91.2<br>(90.7-91.6) | 90.3<br>(89.4-91.1) | 91.0<br>(90.4-91.5) | 91.3<br>(90.8-91.8) | 92.5<br>(91.5-93.4) | <0.0001 |
| Cervix (female) | 74.8<br>(71.3-77.9) | 77.3<br>(73.8-80.4) | 76.4<br>(72.6-79.7) | 77.6<br>(73.8-80.9) | 81.7<br>(78.2-84.8) | 0.0442 | 75.6<br>(72.8-78.1) | 76.2<br>(73.4-78.7) | 79.6<br>(75.1-83.4) | 80.0<br>(77.0-82.6) | 80.1<br>(76.9-83.0) | 84.1<br>(76.3-89.5) | 0.0005 |
| Uterus (female) | 79.8<br>(77.4-82.0) | 82.4<br>(80.4-84.2) | 84.1<br>(82.3-85.8) | 84.1<br>(82.2-85.8) | 82.7<br>(80.9-84.3) | <0.0001 | 82.0<br>(80.5-83.4) | 83.9<br>(82.5-85.2) | 83.8<br>(81.1-86.1) | 83.4<br>(81.7-84.9) | 83.4<br>(81.8-84.9) | 82.9<br>(79.4-85.8) | <0.0001 |
| Ovary (female) | 42.0<br>(38.8-45.3) | 48.3<br>(45.2-51.3) | 45.3<br>(42.3-48.2) | 51.6<br>(48.5-54.6) | 49.7<br>(46.8-52.6) | 0.0001 | 44.9<br>(42.5-47.3) | 46.8<br>(44.6-49.1) | 49.5<br>(45.1-53.8) | 49.6<br>(46.9-52.2) | 47.4<br>(44.6-50.2) | 50.3<br>(44.9-55.4) | <0.0001 |
| Prostate (male) | 96.7<br>(96.1-97.2) | 97.1<br>(96.7-97.5) | 97.5<br>(97-97.8) | 97.8<br>(97.4-98.2) | 98.0<br>(97.7-98.3) | 0.8967 | 96.9<br>(96.6-97.2) | 97.4<br>(97.1-97.7) | 97.6<br>(97.3-97.9) | 97.8<br>(97.5-98.2) | 98.0<br>(97.7-98.3) | 97.7<br>(97.3-98) | 0.0058 |
| Testis (male) | 93.6<br>(90.3-95.8) | 98.3<br>(95.3-99.4) | 98.2<br>(95.9-99.2) | 97.8<br>(95.9-98.8) | 98.5<br>(96.9-99.3) | 0.5546 | 97.6<br>(95.7-98.6) | 97.7<br>(96.3-98.6) | 98.0<br>(96.1-99.0) | 97.3<br>(95.7-98.3) | 98.5<br>(97.3-99.2) | 99.7<br>(91.6-100.0) | 0.4728 |
| Bladder | 75.5<br>(73.2-77.6) | 79.8<br>(78.1-81.4) | 81.9<br>(80.2-83.4) | 80.7<br>(78.9-82.3) | 82.7<br>(81.0-84.3) | 0.0154 | 79.1<br>(77.9-80.3) | 79.9<br>(78.5-81.3) | 81.1<br>(79.5-82.6) | 80.4<br>(78.7-82.1) | 81.9<br>(80.2-83.5) | 85.3<br>(82.6-87.6) | 0.0053 |
| Kidney & renal pelvis | 71.2<br>(68.8-73.4) | 74.9<br>(73.0-76.8) | 77.6<br>(75.7-79.5) | 76.9<br>(75.0-78.8) | 79.0<br>(77.1-80.7) | 0.0017 | 74.5<br>(73.1-75.9) | 76.4<br>(74.8-77.9) | 76.5<br>(74.5-78.4) | 76.8<br>(74.9-78.6) | 78.4<br>(76.6-80.2) | 83.5<br>(80.3-86.2) | <0.0001 |
| Brain & CNS | 17.0<br>(14.5-19.6) | 17.2<br>(14.9-19.6) | 18.5<br>(16.3-20.8) | 15.6<br>(13.5-17.8) | 17.4<br>(15.3-19.5) | <0.0001 | 18.0<br>(16.1-20.1) | 16.6<br>(14.8-18.5) | 15.3<br>(13.0-17.8) | 17.1<br>(15.0-19.3) | 20.1<br>(17.8-22.5) | 22.2<br>(18.2-26.5) | <0.0001 |
| Thyroid | 98.0 | 98.5 | 98.5 | 99.0 | 98.9 | 0.867 | 98.4 | 98.3 | 98.3 | 98.9 | 99.0 | 99.1 | 0.7742 |
|  | (97.1-98.7) | (97.7-99.0) | (97.8-99.0) | (98.3-99.4) | (98.4-99.3) |  | (97.8-98.8) | (97.7-98.7) | (97.4-98.9) | (98.3-99.2) | (98.5-99.3) | (98.1-99.6) |  |
| Hodgkin Lymphoma | 88.3 | 90.8 | 90.9 | 91.0 | 94.9 | 0.3956 | 91.1 | 89.8 | 91.5 | 92.0 | 93.8 | 95.1 | 0.535 |
|  | (84.2-91.4) | (87.7-93.2) | (88.0-93.1) | (88.3-93.2) | (92.7-96.4) |  | (88.9-92.8) | (87.4-91.8) | (88.6-93.6) | (89.5-94.0) | (91.8-95.3) | (91.1-97.3) |  |
| Non-Hodgkin Lymphoma | 67.1 | 72.7 | 73.3 | 73.0 | 78.6 | 0.0012 | 70.4 | 73.3 | 73.2 | 73.6 | 75.8 | 82.1 | <0.0001 |
|  | (64.9-69.1) | (71.0-74.4) | (71.6-74.9) | (71.2-74.6) | (77.0-80.1) |  | (69.1-71.7) | (71.9-74.7) | (71.4-75.0) | (72.0-75.2) | (74.2-77.4) | (79.6-84.3) |  |
| Multiple myeloma | 43.8 | 45.3 | 48.6 | 50.5 | 52.2 | 0.0119 | 45.2 | 45.4 | 48.4 | 48.9 | 51.4 | 58.3 | 0.0006 |
|  | (39.9-47.6) | (42.2-48.4) | (45.3-51.8) | (47.2-53.7) | (49.0-55.3) |  | (42.8-47.6) | (42.6-48.1) | (44.8-51.8) | (45.6-52.1) | (48.3-54.5) | (53.0-63.3) |  |
| Leukemia | 58.2 | 61.0 | 64.8 | 64.9 | 68.7 | 0.0591 | 61.7 | 63.1 | 63.4 | 66.4 | 66.0 | 70.6 | <0.0001 |
|  | (55.4-60.9) | (58.7-63.2) | (62.5-67.0) | (62.6-67.1) | (66.4-70.8) |  | (59.9-63.4) | (61.2-65.0) | (60.9-65.9) | (64.0-68.6) | (63.7-68.2) | (67.0-73.8) |  |

CNS=central nervous system. Age-standardized using the Canadian Cancer Survival Standard weights (individual cancer sites) or the international single standard (all cancers combined).
<sup>a</sup> type III test for interaction effect between the characteristic and cancer status; tests whether there is evidence of heterogeneity in excess cancer mortality rates across groups over the entire 10-year period, not just at 5 years.

### Relative survival compared to life tables

Relative survivals compared to life tables are presented in Table 2 for the overall population, and in Supplementary Table 2 by race and income. Many estimates could not be reported using life tables due to the number of cancer cases and deaths being below the confidentiality disclosure threshold of 5 contributing cases. The relative survival estimates using matched controls were higher than the relative survival estimates using life tables for all cancer sites, with the single exception of brain and central nervous system cancers, which had higher 5- and 10-year relative survival estimates using life table methods (Table 2). While the point estimates changed using different methods, the social gradients in survival by race and income were similar between methods.

### Validation of estimates with historical data

The relative survival estimates using life table methods in CanCHECs 2006 & 2011 were comparable to relative cancer survival statistics based on life table methods for all of Canada during the period of 2006-2017 (Supplementary Methods & Figures). Some cancer sites had slightly lower relative survival estimates in CanCHECs (melanoma, uterine cancer, and leukemias), but the difference was of only 1-2%.

## Discussion

We found significant differences in relative cancer survival by race, immigration status, household income, and education level in Canada for many major cancer sites over 2006-2019. Relative cancer survival was worse in persons with lower household income and lower education levels. Conversely, relative cancer survival was higher in immigrants and in non-White and non-Indigenous racial groups with high proportions of immigrants. Our validation exercise found that relative survival estimates in CanCHECs are comparable to those reported for Canada during the same time period using comparable methods, and consequently our estimates are likely representative of the Canadian population. The cancer survival differences we observed between social groups can be contextualized in terms of the historical progress in cancer control. Relative cancer survival improved by 6% over the 20-year period spanning from 1992/1994 to 2012/2014 in Canada.^2^ The 3-12% survival differences we observed between groups for many cancer sites are therefore analogous to being 10-40 years “behind” in benefiting from improvements in cancer control compared to more advantaged groups.

An important limitation of our analysis is that, despite the combination of two CanCHEC cycles over multiple years of follow-up to increase sample size, there were low event numbers for some cancer sites and demographic groups. This led to issues of power, data confidentiality, and limited ability to examine interactions between the different dimensions of health equity stratifiers affecting survival (intersectionality). While some of the differences in excess cancer mortality rates between groups were not statistically significant, this probably reflects the low number of events in some groups leading to higher uncertainty in estimates. A lack of statistical significance should not be interpreted as indicating an absence of inequalities, but rather a lower power to detect significant differences for some rarer cancers. This issue was particularly important for survival disaggregated by race, as the vast majority of cancer cases were in White persons (88%), due in part to the White population being older and having higher cancer incidence rates than other racial groups in Canada.^28^ This led to lower power to assess clinically meaningful differences by race. While the effects of sex, immigration, household income, and education are also likely to differ by racial group, we were unable to assess these interactions due to low case numbers.

We found that higher household income and education level were consistently associated with improved relative cancer survival across most sites. The majority of previous studies of disparities in cancer survival in Canada have looked at neighborhood-level rather than individual-level measures of income.^7–9^ Our results confirm that these socioeconomic gradients apply at the individual level as well. Canada has a universal health care system funded through taxes, with all citizens and permanent residents having insurance covering most primary and hospital care.^29^ While this partly mitigates inequalities in healthcare access by income, the financial burden of a cancer diagnosis remains unequally distributed as coverage for pharmacy-dispensed prescription drugs and home care is more variable, and cancer patients still face indirect costs such as travel or income loss.^30^ Inequalities in health persist because individuals with more resources such as higher income and education are better able to avail themselves of preventive and therapeutic care to improve their health.^31^ Cancer screening rates display a socioeconomic gradient despite being covered by insurance in Canada.^32,33^ While screening and early diagnosis may contribute to some of the observed differences, previous studies have found that adjusting for stage at diagnosis, however, only partly explain the survival gradient by socioeconomic status.^7^ Differences in treatments received may also contribute to observed differences in survival by socioeconomic status.^8,9^

Studies of immigrants in Canada have documented barriers in access to care such as language and cultural norms.^34^ Immigrants have lower cancer screening participation rates than Canadian-born persons, and are are less likely to have their cancer detected by screening.^35–37^ Nonetheless, our study as well as others consistently find strong evidence of a healthy immigrant effect, where immigrants have generally better cancer outcomes and mortality rates than those born in Canada.^5,28,38,39^ The majority of immigrants to Canada are economic immigrants who are selected based on their ability to contribute to Canada’s economy,^40^ and are required to undergo a medical examination as part of the selection process. This selection process leads to an immigrant population with better average health and performance on a number of health indicators such as fewer chronic conditions as well as lower rates of obesity and smoking than those born in Canada.^41^ The presence of more underlying comorbidities in Canadian-born persons could influence cancer survival through more advanced stages at diagnosis and longer diagnostic intervals, and potentially also differences in tumour biology.^42^

We observed large differences in relative survival by race, though these were not always significant. A large proportion of the individuals from non-White and non-Indigenous racial groups are immigrants,^28^ which may in part explain the higher relative cancer survival across multiple cancer sites compared with White and Indigneous cancer patients, who are predominantly born in Canada. The observed lower cancer survival in Indigenous peoples reflects the enduring legacy of colonialism, which continues to cause barriers in accessing high-quality cancer care for the Indigenous peoples of Canada (the First Nations, Inuit and Métis peoples).^43^ Several prior studies have examined the cancer survival experiences of different Indigenous peoples using a distinctions-based approach.^44–48^

The choice of method to calculate net cancer survival is important when comparing social groups with different background mortality rates, as different methods lead to different results.^49^ In this study, we used relative rather than cause-specific survival to estimate net cancer survival. Cause-specific survival methods use cause-of-death data to estimate net survival from cancer while censoring deaths from other causes (e.g. Kaplan-Meier, Cox models). However, relative survival is generally preferred over cause-specific survival by cancer registries to measure net cancer survival because it avoids errors due to misclassification of the cause of death.^12,13^ Death certificates require a subjective assessment of the cause of death, which may be difficult to resolve in the presence of multiple underlying health conditions. We opted not to use cause-specific survival in part due to differences in the prevalence of comorbities between different groups,^41^ which could lead to differential misclassification of death by the certifier. While relative survival avoids potential misclassification biases from cause of death certification, it can still be a biased estimate of net survival if the comparator is not a good estimate of expected survival in the absence of cancer. Our results suggest that there may be important differences in the background mortality risk of cancer cases compared with general population life tables, and that consequently relative survival compared with life tables likely underestimates net cancer survival, even when using group-specific life tables.

In conclusion, we believe our study fills an important data gap by providing representative national-level estimates of net cancer survival stratified by key individual-level health equity stratifiers for monitoring health equities. We provide estimates using two methods of calculating net survival, including estimates that are comparable with national statistics. Despite universal healthcare access, differences in cancer survival exist by race, immigration status, household income, and education level in Canada, suggesting the root causes of cancer survival inequalities go beyond differences in healthcare access.

## Acknowledgements

Adapted from Statistics Canada, Canadian Census Health and Environment Cohorts 2006 & 2011, 2006 long-form census, 2011 National Household Survey, Canadian Vital Statistics Death Database 2006-2019, and Canadian Cancer Registry 2006-2015. This does not constitute an endorsement by Statistics Canada of this product. The Postal Code^OM^ Conversion File Plus (7D) is based on data licensed from the Canada Post Corporation. The analysis presented in this paper was conducted at the Quebec Interuniversity Centre for Social Statistics (QICSS) which is part of the Canadian Research Data Centre Network (CRDCN). The views expressed in this paper are those of the authors, and not necessarily those of the CRDCN, the QICSS or their partners.

## Data availability

Restrictions apply to the availability of these data, which were used under license by Statistics Canada for this study. Eligible researchers can apply for data access through the Statistics Canada Research Data Centre program (https://www.statcan.gc.ca/en/microdata/data-centres/access). Program code and Poisson model outputs used for the current analyses are available at the Borealis repository at https://doi.org/10.5683/SP3/STER2V

