## Supplementary Methods & Figures for "Inequalities in relative cancer survival by race, immigration status, income, and education for 22 cancer sites in Canada, a cohort study"

### Cancer site classifications

*Supplementary Table 1. ICD-O-3 Topography and histology codes by cancer site.*

| Cancer site | ICD-O-3 Topography | ICD-O-3 Histology (Type) |
|---|---|---|
| All cancers | All sites C00-C80 | All invasive sites |
| Head and neck[a] | C00-C14, C300-C329 | 8000-9049, 9056-9139,9141-9589 |
| Esophagus | C150-C159 | 8000-9049, 9056-9139,9141-9589 |
| Stomach | C160-C169 | 8000-9049, 9056-9139,9141-9589 |
| Colorectal | C180-C189, C260, C199, C209 | 8000-9049, 9056-9139,9141-9589 |
| Liver | C220 | 8000-9049, 9056-9139,9141-9589 |
| Pancreas | C250-C259 | 8000-9049, 9056-9139,9141-9589 |
| Lung & bronchus | C340-C349 | 8000-9049, 9056-9139,9141-9589 |
| Melanoma | C440-C449 | 8720-8790 |
| Breast | C500-C509 | 8000-9049, 9056-9139,9141-9589 |
| Cervix (female) | C530-C539 | 8000-9049, 9056-9139,9141-9589 |
| Uterus (female) | C540-C549, C559 | 8000-9049, 9056-9139,9141-9589 |
| Ovary (female) | C569 | 8000-9049, 9056-9139,9141-9589 |
| Prostate (male) | C619 | 8000-9049, 9056-9139,9141-9589 |
| Testis (male) | C620-C629 | 8000-9049, 9056-9139,9141-9589 |
| Bladder | C670-C679 | 8000-9049, 9056-9139,9141-9589 |
| Kidney & renal pelvis | C649, C659 | 8000-9049, 9056-9139,9141-9589 |
| Brain & CNS | C710-C719 | 8000-9049, 9056-9139,9141-9589 |
| | C710-C719 | 9530 - 9539 |
| | C700-C709, C720-C729 | 8000-9049, 9056-9139,9141-9589 |
| Thyroid | C739 | 8000-9049, 9056-9139,9141-9589 |
| Hodgkin Lymphoma | C000-C809 | 9650-9667 |
| Non-Hodgkin Lymphoma | C000-C809 | 9590-9597, 9670-9719, 9724-9729, 9735, 9737, 9738 |
| | All topographies excluding (C420, C421, C424) | 9811-9818, 9823, 9827, 9837 |
| Multiple myeloma | C000-C809 | 9731-9732, 9734 |
| Leukemia | C000 - C809 | 9826, 9835-9836 |
| | C420, C421, C424 | 9811-9818, 9837 |
| | C420, C421, C424 | 9840, 9861, 9865, 9866, 9867, 9869, 9871-9874, 9895-9897, 9898, 9910, 9911, 9920 |
| | C000 - C809 | 9863, 9875, 9876, 9945, 9946 |
| | C000 - C809 | 9733, 9742, 9800, 9801, 9805, 9806, 9807, 9808, 9809, 9820, 9831, 9832, 9833, 9834, 9860, 9870, 9891, 9930, 9931, 9940, 9948, 9963, 9964 |
| | C420, C421, C424 | 9827 |

CNS=central nervous system; ICD-O-3= International Classification of Diseases for Oncology, 3rd Edition; SEER=Surveillance, Epidemiology, and End Results.

[a] Definition based on the Canadian Cancer Statistics 2021.

### Confidence intervals for relative survival from Poisson models

Cancer relative survival ($RS_t$) at time *t* was calculated by dividing the expected survival for cancer cases ($S_{t,cancer=1}$) by the expected survival for matched controls ($S_{t,cancer=0}$) of the same age, sex, and stratifer level:

$$RS_t = \frac{S_{t,cancer=1}}{S_{t,cancer=0}}$$

The confidence intervals for relative survival estimates ($\widehat{RS}_t$) derived from Poisson regression models were estimated using the log-log transformation of the survival described Kalbfleisch and Prentice:[1]

$$\log\left(-\log\left(\widehat{RS}_t\right)\right)$$

The standard error ($\hat{\tau}(t)$) of this transformation was estimated using the delta method, with the *deltamethod* function of the R *msm* package.[2] The 100(1–α)% confidence interval for relative survival is then given by:

$$\left[\widehat{RS}_t\right]^{\exp\left(z_{\alpha/2}\hat{\tau}(t)\right)} \leq RS_t \leq \left[\widehat{RS}_t\right]^{\exp\left(-z_{\alpha/2}\hat{\tau}(t)\right)}$$

### Life table methods

Life tables compute the death and survival probabilities at different ages for a population, most commonly in 1-year age increments. We used the pooled CanCHECs 2006 and 2011 to estimate cohort-specific life tables stratified by sex, race, and income quintile. For life tables stratified by racial group, the data for Middle Eastern persons was further disaggregated into life tables for Arab and West Asian persons, the data for East Asian persons was further disaggregated into life tables for Chinese, Japanese, and Korean persons, and the data for Southeast Asian persons was further disaggregated into Filipino and other Southeast Asian persons. Follow-up for each age in 1-year increments started on a person's birthday. Each person in the cohort contributed multiple observations for each year they were alive during follow-up (2006-2019).

To estimate yearly mortality hazards, we fitted stratified Cox proportional hazards regression models with all-cause mortality as the outcome, stratified by sex and age for ages 0 to 94 years. Cox models allowed us to include person-time from individuals who did not contribute full years of person-time to each age using the counting process style of input. We included race and income quintile as predictors in the models to calculate race- and income-specific hazards of death. Race and income were included as predictors rather than stratification variables due to issues of data confidentiality and stability of model estimates, as some groups have too few deaths at specific ages to calculate group-stratified hazard functions. The inclusion of these variables as predictors allowed "smoothing" the death hazard function for each group using the assumption of proportional hazards within an age group. Separate models were fit by age group to allow different effects of race and income on mortality in different age groups: 0-14, 15-29, 30-39, 40-49, 50-59, 60-69, 70-79, 80-89, and 90-94 years. The model cumulative hazard functions were used as estimates of mortality rates by age, sex, and race/income. We used the

Fleming-Harrington estimates of the survivor function to estimate probability of death at each age.[3]

For ages 95 and over, mortality rates become too unstable to calculate directly from the data due to low denominator sizes. We used the method employed by Statistics Canada to calculate mortality rates and probabilities for this age group based on projections from a simplified logistic model.[4] The following non-linear model was fit to estimated mortality rates:

$$\mu_x = \frac{\alpha e^{\beta x}}{1 + \alpha e^{\beta x}}$$

Where $\mu_x$ is the mortality rate at age *x,* and $\alpha$ and $\beta$ are fitted parameters determining the shape of the relationship between age and the mortality rate, assumed to be logistic. The model was fitted on the mortality rates estimated for ages <95 years using the Newton method for optimization. The fitted parameters are then used to project expected mortality rates at ages 95 and over. The projected mortality rates at ages 95 and over were then converted into yearly death probabilities ($q_x$) using the actuarial method:

$$q_x = \frac{2\mu_x}{2 + \mu_x}$$

The fitted cohort mortality rates by race are presented in Supplementary Figure 1 below.

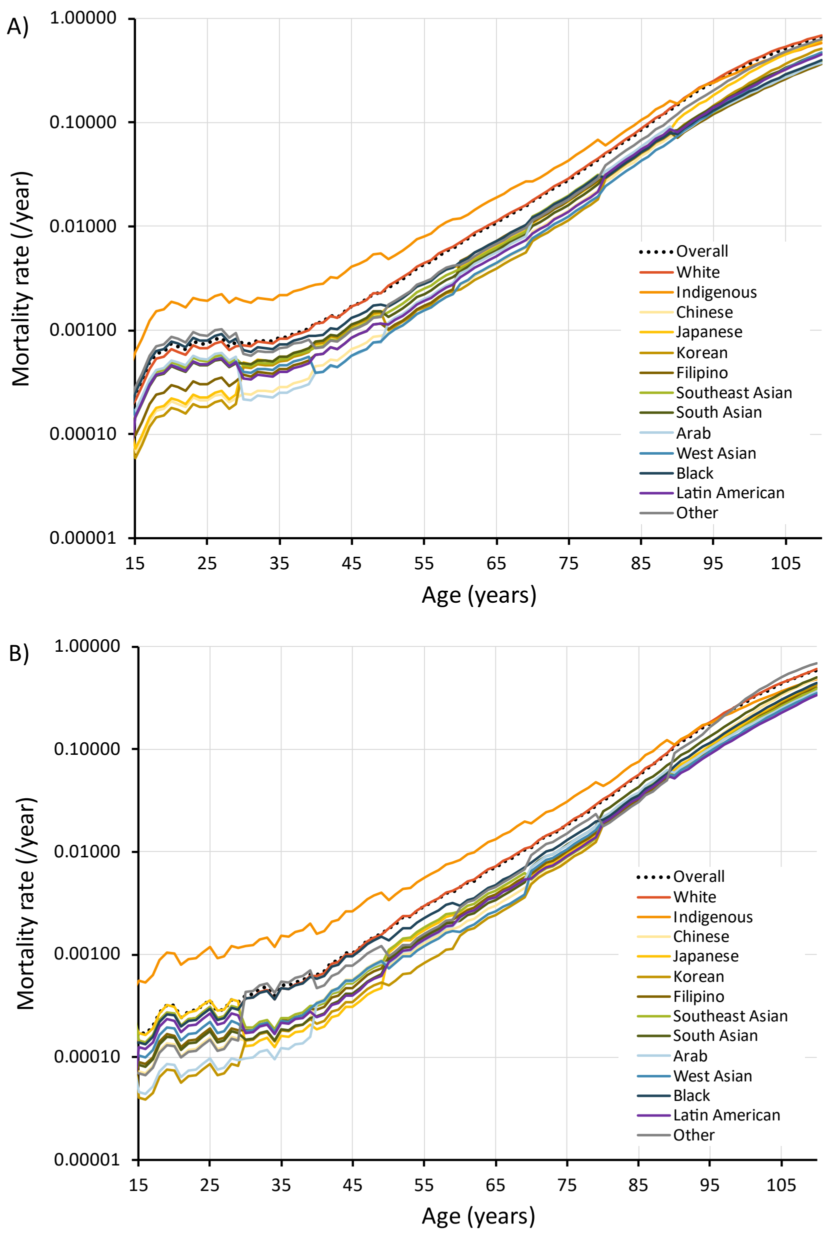


*Supplementary Figure 1. Mortality rates by age and racial group in A) men and B) women in CanCHEC 2006 and 2011. Race-specific probabilities were obtained by fitting age- and sex-stratified Cox regression models with race as a predictor of the hazard, with separate models for ages 15-29, 30-39, 40-49, 50-59, 60-69, 70-79, 80-89, 90-94, and by fitting a simplified logistic model for ages 95+.*

### Relative survival compared with life tables

Period cancer relative survival was calculated by adapting the algorithm developed by Paul Dickman for SAS,[5] which uses the Ederer II method and transformation of the hazard approach. We modified his approach to use the background mortality probabilities estimated from the cohort life tables (described in section above) rather than from external lifetables, and to deal with issues of data sparsity in groups with low case numbers. The mortality hazard in cancer cases was calculated by dividing the number of deaths by the person-time contributed by each cancer case in 1-year intervals up to 10 years. The hazards were then transformed into 1-year interval-specific observed survivals through exponentiation (transformation of the hazard approach). Observed survival was then divided by expected yearly survival from the cohort life tables to obtain relative survival estimates.  In cases where there were no observations contributing to an interval (ex. no cancer cases contributing follow-up time to year 7 after diagnosis in a specific group), the survival probability from the previous interval was carried forward. This assumption is reasonable because issues of data sparsity mostly occurred in the later intervals (5-10 years after diagnosis) when survival probabilities are more similar between intervals. Relative survival was also constrained to be ≤1 in situations where, due to low case numbers, survival was higher in cancer cases than the expected survival based on life tables.

The risk of identifiability is higher when calculating survival using life table methods than with Poisson regression models. With life tables, interval survival is calculated using a direct transformation of the number of events, so large drops in the survival curve are more easily identified as being due to one or a few deaths within an interval when there are small denominators; with Poisson models, the additional parametric assumptions mask the contribution of individual events. Estimates where fewer than 5 deaths contributed to cumulative relative survival in a group were suppressed when using life table methods in accordance with confidentiality disclosure rules. This was more likely to occur for cancer sites with low incidence and mortality rates (e.g. testicular cancer) and for groups with small denominator sizes (e.g. Latin American cancer cases). Because cumulative survival closer to diagnosis is based on fewer events (deaths), 1-year survival estimates were also less likely to meet confidentiality disclosure thresholds than 5- or 10-year survival estimates due to lower cumulative event numbers.

### Comparison of CanCHECs with historical data

Relative survival estimates using life table methods for the overall CanCHECs 2006 & 2011 are shown in Supplementary Figures 2-4, and compared with estimates from the Canadian Cancer Statistics for the periods of 2006-2008 and 2015-2017. Most confidence intervals overlap.

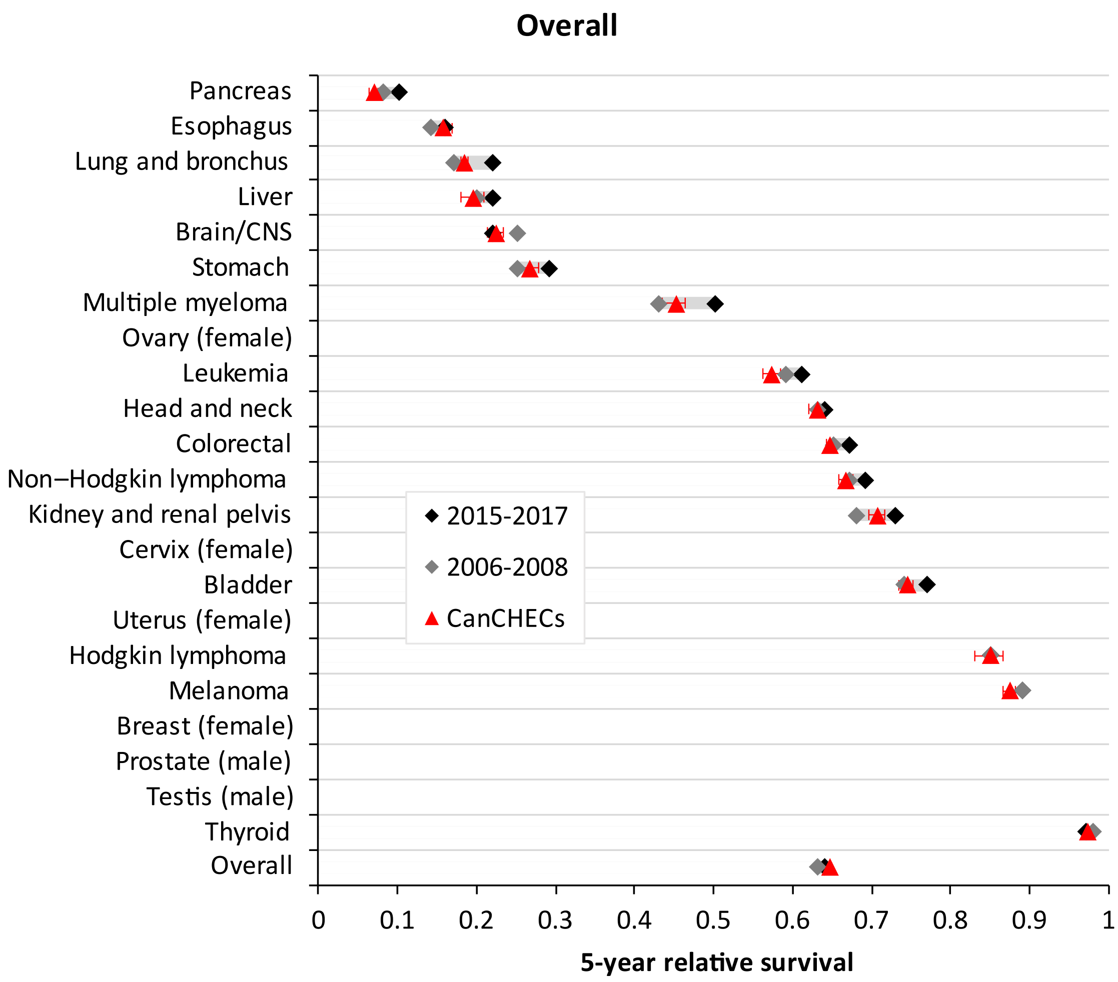


*Supplementary Figure 2. Comparison of historical relative survival in Canada with estimates of period relative survival for CanCHECs 2006 & 2011, both sexes combined, ages 15-99. Note that due to the new definition for head & neck cancers in 2021, we used oral cancer survival as a proxy for 2006-2008. Data source: Canadian Cancer Statistics 2013 & 2021.*[6,7]

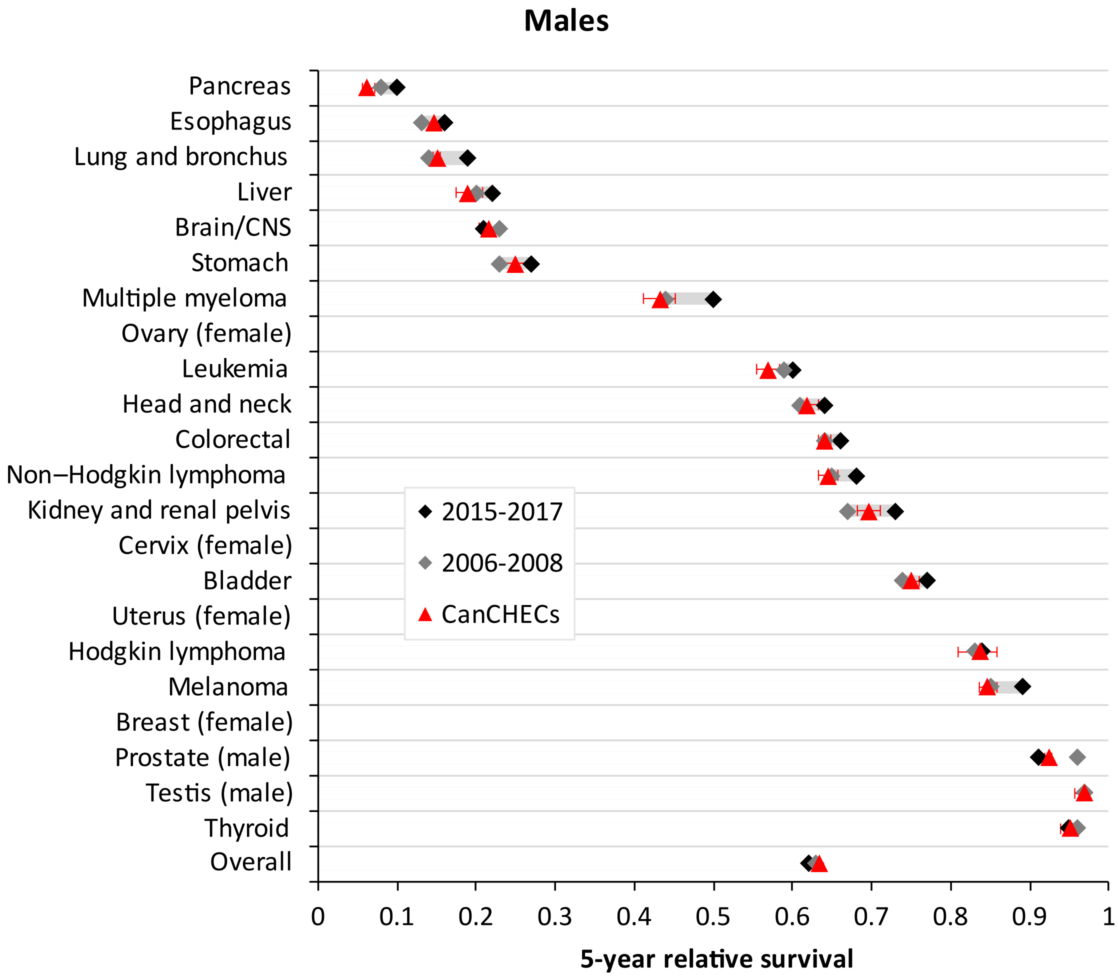


*Supplementary Figure 3. Comparison of historical relative survival in Canada with estimates of period relative survival for CanCHECs 2006 & 2011, males ages 15-99. Note that due to the new definition for head & neck cancers in 2021, we used oral cancer survival as a proxy for 2006-2008. Data source: Canadian Cancer Statistics 2013 & 2021.*[6,7]

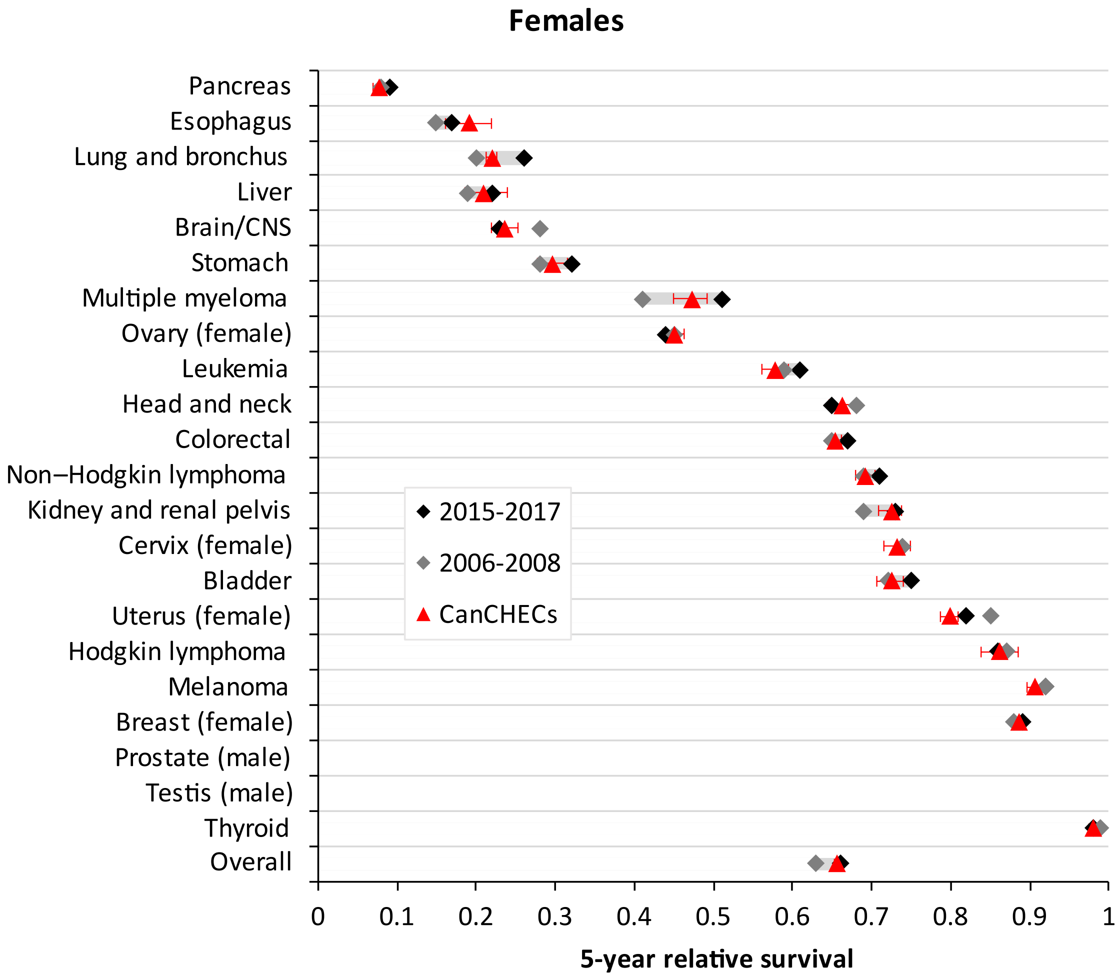


*Supplementary Figure 4. Comparison of historical relative survival in Canada with estimates of period relative survival for CanCHECs 2006 & 2011, females ages 15-99. Note that due to the new definition for head & neck cancers in 2021, we used oral cancer survival as a proxy for 2006-2008. Data source: Canadian Cancer Statistics 2013 & 2021.*[6,7]
